# *“Someone who hates themselves doesn’t come for their drugs”:* Experiences of Mental Health Along the HIV Care Continuum in South-Central, Uganda

**DOI:** 10.1101/2023.08.24.23294215

**Authors:** Nora S. West, William Ddaaki, Sarah M. Murray, Neema Nakyanjo, Dauda Isabirye, Rosette Nakubulwa, Fred Nalugoda, Pamela J. Surkan, Heidi E. Hutton, Caitlin E. Kennedy

**Affiliations:** Division of Pulmonary and Critical Care Medicine, University of California San Francisco, San Francisco CA USA; Rakai Health Sciences Program, Kalisizo, Uganda; Department of Mental Health, Johns Hopkins Bloomberg School of Public Health, Baltimore, MD, USA; Department of International Health, Social and Behavioral Interventions Program, Johns Hopkins Bloomberg School of Public Health, Baltimore, MD, USA; Division of Psychiatry and Behavioral Sciences, Johns Hopkins School of Medicine, Baltimore, MD, USA

**Keywords:** HIV, mental health, qualitative research, continuum of care, Africa

## Abstract

**Introduction:** Poor mental health occurs more frequently among people living with HIV. Understanding what mental health problems occur and at what point during the continuum of HIV care is critical to ensure these problems are identified and appropriately addressed. We explored how mental health is experienced along the HIV care continuum in Rakai, Uganda.

**Methods:** We conducted qualitative semi-structured in-depth interviews with 20 adults living with HIV and 10 health workers from March to December 2020. Interviews followed a timeline approach. Responses were analyzed using content analysis.

**Results:** At the time of HIV diagnosis, nearly all participants described a range of strong emotions, including shock, fear and intense worry. Most participants described continued fear and intense worry leading up to, and at the time of, ART initiation. However, they said these emotions often subside after ART is initiated and viral suppression is achieved. Across interviews and at multiple points of the continuum, participants discussed how fear and worry led individuals to be “thinking too much” or be in “deep thoughts” and experience self-hatred. Individuals who stopped taking ART were thought to have more severe mental health problems (“madness”, psychosis, suicidality). Participants were divided about the mental health of persons who returned to care after disengagement.

**Conclusion:** In this setting, mental health problems experienced by people living with HIV are dynamic across the care continuum. With expanded HIV testing campaigns and Universal Test and Treat policies, targeted interventions for psychosocial support at the time of testing and ART initiation remain critical.

## INTRODUCTION

HIV persists as a leading cause of death in sub-Saharan Africa (SSA) [1], highlighting a continued urgent need to address the factors that contribute to poor outcomes for people living with HIV [2]. HIV prevalence in Uganda is 5.9% nationally [3] but significantly higher in the predominantly rural Rakai region where median prevalence ranges from 14% in agrarian communities to 42% in fishing communities [4, 5]. Explanations for this markedly elevated HIV prevalence are multifactorial, and include high levels of in and out cross-community migration in the Rakai region, a historical lack of targeted HIV testing and treatment services and service uptake in the region, and the presence of fishing and trading communities where individuals with high levels of HIV-related sexual risk behaviors are more likely to reside [4, 6]. There remains a pressing need to both effectively manage the health of people already living with HIV and prevent new infections in the Rakai setting.

The HIV care continuum is an HIV care and treatment framework that depicts the often bi-[7]directional spectrum from HIV diagnosis, linkage to care, retention in care, and viral suppression [8]. In the modern treatment era of Universal Test and Treat (UTT) all newly HIV diagnosed individuals are eligible for ART regardless of CD4 count. Herce et al. have further conceptualized the linkage to care step as a multifaceted stage of the continuum, inclusive of post-test counselling, care transfer, clinical assessment, ART initiation, counselling and support, and first follow-up – suggesting it is not as brief or simple a step as it might appear [9].

Evidence from SSA demonstrates that poor mental health, inclusive of symptoms or diagnosis of a mental disorder or disorders, occurs more frequently among people living with HIV than the general population [10–13]. In addition to being common, poor mental health and psychological distress among people living with HIV often go unrecognized and have deleterious impacts on further HIV transmission and treatment outcomes [14]. Beyond the impact on general wellbeing for people living with HIV, poor mental health and HIV-related stigma are linked to engagement in risk behaviors including delayed care seeking, decreased medication adherence, and poor retention in care among people living with HIV in low- and middle-income countries (LMICs) [10, 11, 13, 15–22]. Despite this important evidence, a clear picture of both the types of poor mental health that occur and the mental health trajectories within and across the continuum in the modern treatment era (i.e. since the adoption of UTT policies) is lacking. UTT critically ensures that all people living with HIV can access lifesaving treatment. However, there is evidence that individuals initiated immediately after HIV diagnosis (i.e. same day) may have worse treatment outcomes than individuals who are initiated slightly later (i.e. within 2 weeks following diagnosis), suggesting there may be nuance in the time and support individuals need following HIV diagnosis [23, 24]. In the SSA setting of readily available and rapidly initiated ART, the impact of mental health on achieving favorable HIV treatment outcomes is still largely unknown. Further, studies from SSA in the UTT-era typically explore mental health at just one or two steps of the continuum, and often focus only on symptoms of a singular disorder (primarily depression) [25]. This evidence gap needs to be addressed.

Additionally, understanding the timing and spectrum of mental health problems, and how they may wax and wane along the HIV care continuum, is important for developing tailored and timely interventions. This comprehensive understanding of mental health along the HIV care continuum may be critical to achieving optimal mental health and HIV treatment outcomes and is absent in the literature from SSA. We used qualitative research to explore how mental health is experienced along the HIV care continuum in Rakai, Uganda.

## MATERIALS AND METHODS

### Theoretical Frameworks

This research was influenced by the ethnomedical theoretical perspective. Central to this theory is that health, illness, and individuals’ perceptions of health and illness are embedded in social and cultural contexts [26]. This theoretical perspective guides the exploration of local experiences, terms, practices, and models related to mental health, as well as the pertinent connections between mind and body. Furthermore, it emphasizes that insights gained from research employing this theoretical framework can enhance future health research and programming [26]. In addition, the design of the interviews drew upon journey mapping, a health research approach used to linearly describe an individual’s path, often a person with an illness, to elucidate meaningful patterns along the path – in this case, along the HIV care continuum [27].

### Study Setting and Population

From March 2020 to December 2020, we conducted semi-structured in-depth interviews (IDIs) with people living with HIV and health workers in Rakai, south-central Uganda. Adults living with HIV (ages 18-49) were recruited from participants in recent rounds of the Rakai Community Cohort Study (RCCS), an open ongoing population-based cohort study across 40 predominantly rural communities in the Rakai region, who had previously agreed to be recontacted for future studies [4]. Briefly, the 40 study communities, selected to be representative of the Rakai region, undergo a census roughly ever 1-2 years where all households within the community clusters are systematically approached and enumerated [4]. Following the census, the RCCS enrolls eligible participants in central community locations, with follow-up visits to households that do not present in attempts to enroll all households in the enumerated community cluster. Participation in the RCCS is high, with up to 99% of eligible individuals participating in previous data collection rounds [4]. Full details of the RCCS design and methods are published elsewhere [4, 28].

We purposively sampled participants for our study to achieve balance in time since HIV diagnosis, based on available data from the RCCS (diagnosis ≤18 months or >18 months ago), and gender. Sampling on these characteristics was chosen to allow for the exploration of similarities and differences in psychological distress across these different dimensions and at different time points in the HIV care continuum. Health workers who regularly provide care to people living with HIV and who were employed by any of the three Rakai Health Sciences Program (RHSP)-affiliated clinics were also eligible to participate in the study. Health workers were selected based on recommendations from RHSP staff with a focus on health workers knowledgeable about the mental health and psychosocial needs of people living with HIV.

### Procedures

Among individuals recruited for the study, two people living with HIV declined participation due to inability to commit the time needed for the interview. The majority of IDIs were conducted in Luganda by experienced RHSP qualitative data collectors, including authors R.N. and D.I.. Four IDIs with health workers were conducted in English by author N.S.W.. All participants provided informed consent. Four interviews were conducted in-person in March 2020 among people living with HIV, while all subsequent interviews were carried out via telephone due to COVID-19 pandemic lockdown restrictions. Participants who completed in-person interviews provided written consent while participants who completed telephone interviews provided oral consent. All IDIs were completed at a single timepoint and lasted from 45-90 minutes.

Semi-structured interview guides (Appendix 1) were used and followed a timeline representing the HIV care continuum, starting from before HIV diagnosis and moving through retention in care. Participants were asked about the experiences of people living with HIV in their community, and not about their personal experiences. Specifically, they were asked about the problems related to emotions, feelings, thinking, or mental health that people who have HIV might experience at each step along the timeline. Interview guide questions were developed by the research team based on research objectives and theory and adapted from pre-existing questions piloted and utilized in other studies when applicable. Interview guide questions were designed to elicit descriptions of mental health and/or distress along the HIV care continuum without presupposition of how this might manifest or be named by participants. Thus, interview guides did not ask about specific mental disorders or types of psychological distress but used open ended questions to facilitate sharing of this information by participants. Separate questions were used to ask participants about the drivers for the type or types of poor mental or psychological distress health that participants described. Questions about the impact of the COVID-19 pandemic on the psychological wellbeing of people living with HIV were asked for all participants who had telephone interviews conducted. These methods and findings have been reported elsewhere [29]. Interview guides were developed in English and translated to Luganda by bi-lingual research team members through an initial process of translation and back translation, followed by team-based review and discussion before arriving at final Luganda and English versions of the interview guides. All interviews were audio recorded. All interviews were transcribed, and interviews conducted in Luganda were translated into English. Maximum variation sampling for qualitative research, where sample size is determined by number of factors sampled and planned sample homogeneity considered, was used to determine planned samples sizes for each participant group [30]. The principle of saturation, when new or additional data does not yield new information, was used to arrive at the final sample size [31]. During data collection, study investigators met weekly with the data collection team to discuss interview progress and content, review debrief forms and notes, and available interview transcripts to assess saturation throughout data collection.

### Analysis

Content analysis was used as the analytical approach [32]. A codebook was initially developed by N.S.W. based on the interview guide and further refined through reading and re-reading of the transcripts, initial coding, and analytic memoing. Codes and interpretations were shared with the data collection team and study investigators throughout the analysis process for input, feedback, and refinement. N.S.W. coded all transcripts using the final version of the codebook. Issues discussed in the interviews were grouped by step of the HIV care continuum and further categorized by specific stressors or mental health problems at each step. Findings were examined and compared based on sampling stratifications and participant category for the exploration of similarities and differences across these different dimensions.

The study was approved by the Institutional Review Boards of the Johns Hopkins Bloomberg School of Public Health, The Research and Ethics Committee of the Ugandan Virus Research Institute, and the Uganda National Council on Science and Technology.

## RESULTS

The sample included 20 people living with HIV (n=11 women, n=9 men; n=11 diagnosed ≤18 months ago, n=9 diagnosed >18 months ago) and 10 health workers (n=5 nurses, n=3 HIV counsellors, n=2 peer health workers). Moving along the HIV care continuum, participants described factors shaping psychological distress, mental health and resulting mental health outcomes at the different stages of the continuum. Findings are presented by stages of HIV care continuum with exemplary quotes. Figure 1 shows an overall summary of findings.

**Figure 1.**
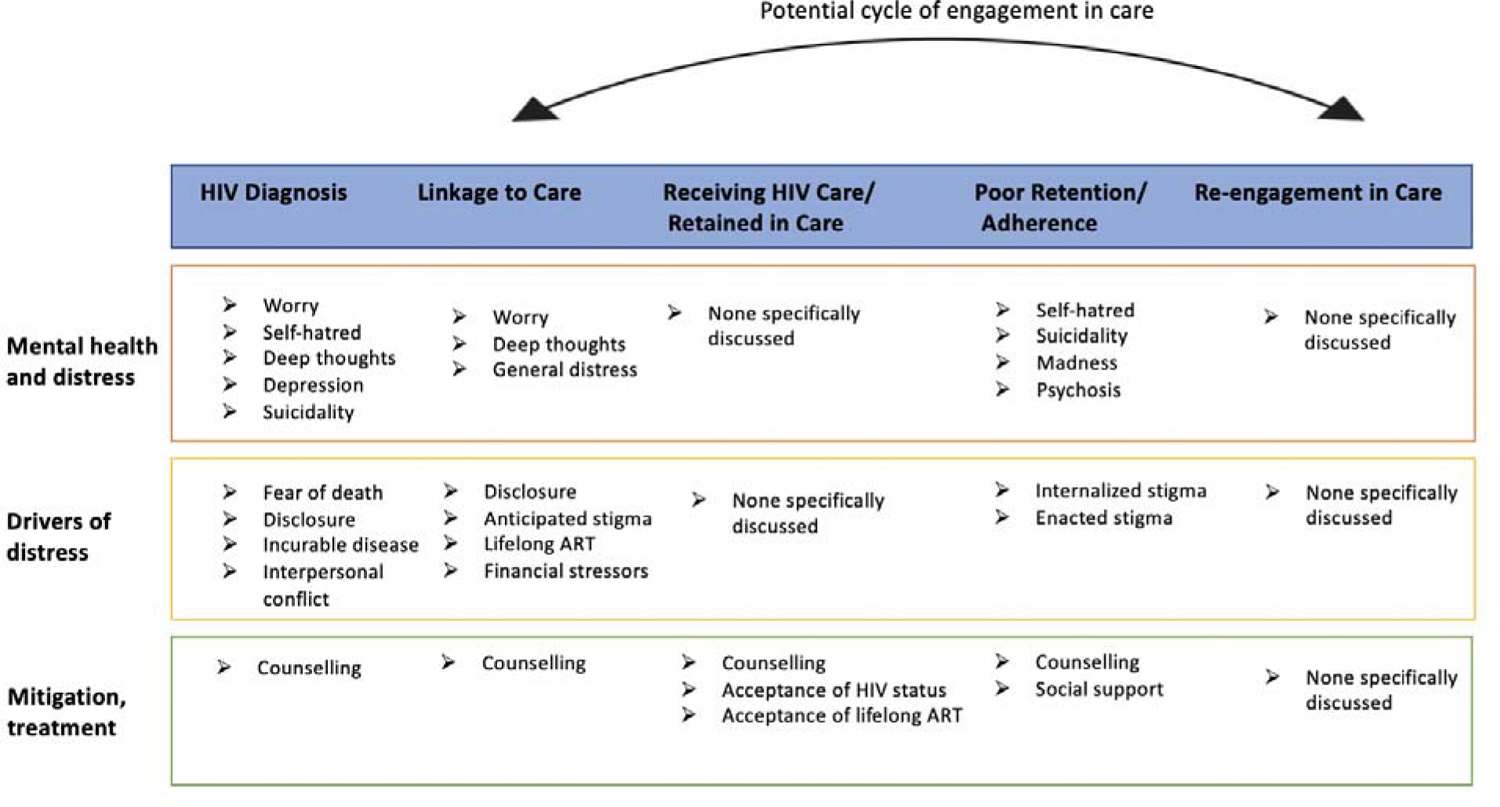
Descriptors of Mental Health and Distress, Drivers of Distress, and Mitigation or Treatment Approaches.

### HIV diagnosis

Most participants felt that prior to HIV testing, people living with HIV did not experience mental health problems, although the stressors of daily life regardless of HIV status were always present. At the time of diagnosis, all but one participant said that people living with HIV experience a range of strong emotions with potential mental health impacts. Prominent throughout most interviews at the time of HIV diagnosis was the fear of death and the idea that HIV is a death sentence, which in turn could cause mental health issues, as described by a peer health worker:

> If you had just provided HIV positive test results to the client, this person asks him or herself what to do next, and this causes depression. It is not easy to accept it…secondly, a person starts thinking about his or her future life, and how to stay alive. Thirdly, he or she will say ‘nkomye’ [this is the end of my life], and this makes an individual get worried. This person assumes that being HIV positive marks the end of life, and no more chances to live. As the client continues to cry, he or she says, ‘this is the end of my life, how should I survive.’

Fear of death, described by most participants, was sometimes defined by causing someone to experience *okwenyamira* (translated as “deep thoughts” or “many thoughts” roughly defined as ongoing unsettling or unending thinking). Other feelings of fear could be driven by concerns about HIV disclosure and worry about not being able to care for children/family, as described by a man diagnosed with HIV ≤18 months ago: “*If someone has tested and found HIV positive…you get deep thoughts knowing that it is the end of your life*.” These concerns, including having to disclose one’s HIV status led to immediate distress over the idea of having to initiate ART and the tradeoffs between initiating ART or not.

About half of participants said that self-hatred and suicidal thoughts could follow an HIV diagnosis, as many newly diagnosed individuals were reported to grapple with the idea of a lifelong disease. *“After some people are diagnosed with HIV, they start hating themselves. He/she might be there and starts thinking of taking poison and dying. After all, I will not be cured.”* (Female, diagnosed >18 months ago). Many participants, particularly those who had been diagnosed In the past 18 months, said that worry and anxiety over having to either disclose to others or conceal their HIV status was prominent at the time of diagnosis.

### Linkage to Care

The majority of participants talked about how the time leading up to or at ART initiation was a period of significant distress. Reasons for this were multi-faceted, with fear about inadvertent status disclosure and stigma as prominent drivers of worry (*okweraliikirira*) and “deep thoughts” (*okwenyamira*). Anticipated stigma from family and community members because of recent HIV diagnosis in advance of ART initiation was more commonly talked about by participants more recently diagnosed with HIV, and enacted stigma was only raised by this group, as described by a woman, diagnosed with HIV ≤18 months ago:

> A family member can see you taking HIV medicine and begins telling people that “so and so is taking ARVs”. When you get to know that -- you begin thinking that wherever I go people talk/gossip about me having HIV. You begin feeling uncomfortable or unstable…they make them feel totally uncomfortable. This makes you be in deep thoughts.

Fear of side effects, having to take a daily pill, and worry about needing money to eat well while taking ART and to get to the clinic were other commonly mentioned by many participants as causes of distress at the time of ART initiation. Economic difficulties were discussed in general as a source of stress in all people’s lives, but compounded for people living with HIV because of having to access care and eat well when on ART.

While a few participants felt that people living with HIV may not experience any mental health issues in the time leading up to ART initiation, many described post-test counselling or on-going counselling leading up to or at the time of ART initiation as an important way of alleviating emotional distress following HIV diagnosis. *“If it is not for intensive counseling that organizations and government or the ministry have put in place to be given to people [recently diagnosed with HIV], maybe these tendencies of suicide would be many.”* (HIV Counsellor)

### Receiving HIV Care, Retention in Care, and Viral Suppression

Overall nearly all participants felt that individuals who were receiving HIV care, adherent, and retained in care did not experience many mental health problems in large part because they had been able to receive counselling, accept their diagnosis, and come to terms with lifelong ART. When discussing mental health-related reasons that people living with HIV might stop taking ART or have trouble adhering, a number of participants said that self-hatred could be a catalyst: *“If you hate yourself* [okwekyawa]*, even if you fetch the drugs, you are not going to take it because you are fed up. Someone who hates themselves doesn’t come for their drugs.”* (Female, diagnosed >18 months ago). A number of participants talked about discontinuing treatment as a death wish or akin to being suicidal. Both health workers and people living with HIV said that the downstream impact of discontinuing ART was madness or psychosis, sometimes attributed to untreated HIV-infection. *“If a person stops HIV care and treatment completely, it results in ‘*okufa*’ [death] or ‘*okwagala okwetta*’ [suicide attempt]. He/she first experiences ‘*kulaluka’ *[madness] and then later dies. You cannot stop HIV care and treatment knowing that you can only live by ARVs. ARVs are your life.”* (Female, diagnosed >18 months ago). Most participants said that people living with HIV who were experiencing mental distress and struggling with adherence and retention as a result needed counselling at the clinic and/or social support from family members and others living with HIV to mitigate these problems and described how through counselling and support people living with HIV could become “strong” and overcome distress.

When talking about people living with HIV who had been re-engaged in care, participants had mixed opinions about what mental health problems might be experienced. Some participants felt that once someone had re-engaged, they were not experiencing problems or had overcome any mental health issues that would have been a barrier to re-engagement. A nurse said, *“Indeed, I do not know because I would not expect the client who had re-started taking ART to face any [emotional or mental health] problem. The client has already explained that he or she is now re-starting the HIV treatment, I do not see any likely problem this client would face.”* However, some participants felt that the problems experienced at the HIV diagnosis or treatment initiation timepoints might reoccur.

## DISCUSSION

This qualitative study explored how people living with HIV in South-Central, Uganda experience psychological distress and poor mental health as they encounter different steps and progress along the HIV care continuum. Overall, participants felt that poor mental health was rare prior to testing and at the time of HIV testing participants described varying types of distress. Initiating ART caused worry, fear, and “deep thoughts”, with anticipated HIV-related stigma as a prominent driver. Those who struggled with adherence or dropped out of care were thought to experience self-hatred and be potentially suicidal, with possible long-term impacts of “madness” or psychosis. When people living with HIV re-engaged in care, participant responses were mixed as to whether the cycle of stressors along the continuum and their impacts on mental health would repeat upon re-starting ART.

The 95-95-95 goals have importantly led to the scale-up of HIV testing and adoption of differentiated approaches such as client or provider-initiated, community-based, or self-testing.[33] However, the relationship between mental health and HIV diagnosis is complex, with little empirical data from SSA to demonstrate whether poor mental health precedes HIV infection, leads to a delay in HIV testing, or is the result of an HIV diagnosis.[13] Our findings suggest that HIV diagnosis can prompt psychological distress that may lead to poor mental health, and anticipation of a positive HIV diagnosis or fear of diagnosis may also drive distress prior to engagement in the care continuum. In our study, post-test counselling was felt to be one important way to alleviate distress following HIV-diagnosis. Client-centered post-test counselling that is tailored to the needs of a newly diagnosed individual has been recommended by the WHO, although guidance on post-test counselling content is generally focused on preparation for ART and linkage to care,[33] leaving a critical gap for addressing the potential immediate need for psychosocial support following a new HIV diagnosis which may further complicate an individual progressing to the linkage to care step. Neither recent WHO or Ugandan post-test counselling guidelines emphasize psychosocial support or assessment for mental health problems immediately following HIV diagnosis as part of post-test counselling.[33, 34] Incorporating basic psychosocial support into post-test counselling following HIV diagnosis may help mitigate distress and/or prevent worsening distress that could lead to mental health issues. Additionally, screening for poor mental health at other steps of the continuum (such as linkage-to-care) may identify individuals to prevent development of mental disorder.

Prompt initiation of ART is important to ensure the health and wellbeing of people living with HIV and prevent onward transmission, but our findings suggest that rapid ART initiation may be accompanied by multifaceted psychological stressors that are experienced during linkage to care, supporting the idea that attention to mental health for those in need of additional support may be critical to their progression along the continuum.[14] In the era of UTT, more intensive intervention for individuals experiencing poor mental health and distress around ART initiation may be warranted to aid in progression through this critical timepoint in HIV care. Case management, often peer-led, that includes psychosocial support has been used as an approach to improve linkage to care and increase the likelihood of ART initiation.[35] Additionally, our findings demonstrate that anticipated or enacted stigma may contribute to the likelihood of psychological distress at the linkage to care step. This is in line with a substantial body of literature demonstrating associations between HIV-related stigma (anticipated, enacted, internalized) and poor mental health for people living with HIV.[14, 22] In our study anticipated and enacted stigma were raised primarily by participants who were more recently diagnosed with HIV (≤18 months ago), suggesting that individuals may be actively grappling with concerns around stigma and disclosure based on the recency of their diagnosis and therefore find these issues to be critical. Given the influential role that HIV-related stigma plays both for mental health and in HIV care and treatment outcomes for people living with HIV,[22] and that participants in this study who highlighted stigma the most were more recently diagnosed, attention to stigma early on in the continuum of care may be warranted.

The relationship between common mental disorders or symptoms of common mental disorders and adherence to ART and/or engagement in HIV care has been well studied.[14, 36–38] Our findings further support that psychological distress and poor mental health may be key contributing factors for individuals who struggle with adherence and retention in care.

Participants in our study were divided on whether re-engagement in care meant an individual would not experience or was no longer experiencing poor mental health or distress, or whether having to progress again through the HIV care continuum would cause someone to experience psychological distress and potentially poor mental health outcomes all over again. Research on mental health and re-engagement in HIV care is scant. A US-based study found that people living with HIV who had a mental disorder were less likely return to care after a single HIV visit followed by a subsequent 6-month gap in care.[39] A study among people living with HIV re-engaging in care in Argentina found that 21% reported suicidal ideation in the past week.[40] We were unable to identify any literature from sub-Saharan Africa examining mental health among individuals who return to care, but our findings suggest that this may be a time when mental health stressors are renewed and psychosocial support is particularly needed. Individuals re-engaging in care may benefit from more intensive mental health screening, support, and follow-up.

Suicidal ideation and/or attempt was discussed frequently by our participants – particularly in the HIV diagnosis and linkage to care steps, and for people who had disengaged with care. A systematic review and meta-analysis of people living with HIV in Africa found prevalence of suicidal ideation to be 21.7% and a prevalence of suicide attempt of 11.1%.[41] This review did not examine timing of ideation and/or attempt, but our findings suggest these may occur across the continuum of care. Depression is associated with suicidality among people living with HIV in Africa.[42–45] While the Ugandan HIV guidelines state that annual screening for depression should occur within the healthcare setting for people living with HIV,[46] the reality of limited resources both to conduct screening and to refer people living with HIV in need of further support are likely barriers to the implementation of these guidelines. Furthermore, health workers may not be adequately trained to manage comments about suicide attempts or ideation and address them in the context of potential underlying mental disorders.

This study is not without limitations. First, participants were not asked about their personal experiences or sampled based on mental health status or level of engagement with other people living with HIV, but rather asked to reflect generally on the experiences of people living with HIV in their communities. This limits our ability to explore lived personal experiences and trajectories along the care continuum; however, may have allowed people to speak more freely about stigmatized issues. Despite this limitation, this approach does allow for an exploration of the overall perceptions of what participants see as the issues experienced in their communities as opposed to simply an individual experience. Furthermore, a number of participants chose to share their personal experiences when providing examples of mental health challenges along the HIV care continuum. Second, data were collected in predominantly rural areas which may limit transferability of the findings to urban settings. Third, interviews were restricted to individuals who agreed to be contacted for future research, and the majority were conducted via telephone, which may have limited who was able to participate based on prior consent and telephone access. However, the RCCS from which participants were drawn has procedures in place to obtain reliable telephone contact information from all cohort participants and the majority of RCCS participants agree to be contacted for studies, which may have mostly mitigated these concerns in our sample.

## CONCLUSION

These findings suggest mental health problems experienced by people living with HIV are not necessarily uniform along the HIV care continuum and may require nuanced intervention approaches. With expanded HIV testing campaigns and UTT policies, tailored and targeted interventions for psychosocial support remain critical. Given the important and persistent role of anticipated or enacted stigma as a driver of poor mental health found in this study and throughout the literature,[16] continued attention is warranted on the implementation of evidence-based approaches to decrease stigma at the community and individual levels. Finally, more research is needed to understand the mental health needs of individuals who are returning to HIV care. Careful consideration of both the catalysts for psychological distress and resulting poor mental health at each step along the HIV care continuum are critical to ensuring the best possible outcomes for people living with HIV throughout their HIV care and treatment.

## Data Availability

Data cannot be shared publicly because of content in the informed consent that states no unauthorized individuals outside of the research study will have access to participants data. Reasonable requests for data will be considered by the corresponding author on an case by case basis.

## Acknowledgements

The authors wish to thank the participants who provided their insights and time to this study. We also thank the broader Rakai Health Sciences Program Social and Behavioral Studies team who supported this work.

